# The effect of playing career on chronic neurophysiological changes in retired male football players. An exploratory study using transcranial magnetic stimulation

**DOI:** 10.1101/2024.05.28.24308010

**Authors:** Alan J Pearce, Jamie Tallent, Ashlyn K Frazer, Billymo Rist, Dawson J Kidgell

**Affiliations:** School of Allied Health, Human Services and Sport, La Trobe University, Melbourne, Australia, 3086; School of Sport, Rehabilitation and Exercise Sciences, University of Essex, Essex, UK, CO4 3SQ; Faculty of Medicine Nursing and Health Science, Monash University, Melbourne, Australia, 3800

**Keywords:** Ageing athletes, professional football, neurotrauma, transcranial magnetic stimulation, pathophysiology, concussion, traumatic encephalopathy syndrome

## Abstract

**Aim:** Repetitive head impact exposure, from contact and collision sports, are increasingly being attributed to increased risk of neurodegenerative disease in aging athletes. This exploratory study investigated the association of playing career in retired professional contact sport athletes with cortical neurophysiology via transcranial magnetic stimulation (TMS).

**Methods:** Male athletes between the ages of 28-68 years (n=113; mean age [SD] 48.8 [9.7]) who had been retired from professional sport for a minimum of five years were recruited. Cortical excitability was measured using single pulse TMS for motor evoked potentials and paired pulse transcranial magnetic stimulation short-interval intracortical inhibition and long-interval intracortical inhibition. Associations were assessed between transcranial magnetic stimulation measures and concussion history, clinical symptom scores, total career length (including junior to complete retirement), and professional career length (elite competition only).

**Results:** Correlations showed significant associations between motor evoked potentials and clinical symptom reporting (*rho*: -0.21 – -0.38; *P*<0.01); and motor evoked potentials and short-interval intracortical inhibition with total career length (*rho*: 0.26 – -0.33; *P*<0.01). No significant correlations were observed between single and paired-pulse transcranial magnetic stimulation and professional career length (*rho*: 0.16 – -0.15), nor the number of concussions (*rho*: 0.17 – -0.17).

**Conclusions:** This study is the first to report pathophysiological outcomes in a cohort of retired professional athletes associated with total career exposure, rather than professional career exposure or concussion history. TMS assessment could be considered a viable biomarker in future studies of retired athletes suspected with traumatic encephalopathy syndrome.

---

Cumulative exposure to repetitive concussion and sub-concussion impacts are increasingly being attributed to abnormal ageing (1), neurological (2), and cognitive impairments (3), and increased risks of neurodegenerative disease (2, 4, 5). Currently a definitive diagnosis of neurodegenerative disease, particularly chronic traumatic encephalopathy (CTE), can only be confirmed via neuropathology (6, 7). However, in understanding the progression of CTE, descriptions of the clinical characteristics of CTE include reports of non-specific symptoms, cognitive, behavioural, mood and other psychiatric features, and motor impairments. These characteristics have contributed to the formation of a criteria for traumatic encephalopathy syndrome (TES) (6).

The TES clinical diagnostic criteria, which is currently in progress, acknowledges the need to include biomarkers (6). While specific biomarkers have not yet been accepted, there is some promise from studies utilising transcranial magnetic stimulation (TMS) (8–10). First reported in 1985 (11, 12), TMS is a well-established technique that can non-invasively assess cortical physiology in health and disease (13). In abnormal ageing and neurology, TMS has shown to have clinical diagnostic utility (14), including neurodegenerative diseases. Particularly in dementia, the loss of synaptic density and abnormalities in neurotransmission have been detected using TMS in Alzheimer’s disease (AD) compared to healthy older adults (15). Consequently, TMS has been touted as a potential prodromal biomarker for diagnosis of AD or frontotemporal dementia (15–18), and may have potential as part of the overall criteria for TES.

In understanding long term concerns from repetitive neurotrauma, previous TMS studies in retired athletes have specifically focussed on concussion history, for example mean number of concussions, and the time since last reported concussion (8, 9, 19, 20). These studies showed changes in cortical inhibition compared to age-matched controls but did not consider playing career length exposure. Similarly, a recent TMS study in contact sport athletes showed decreased cortical inhibition verses age-matched controls that worsened with ageing (10). However, that study did not consider effects of playing career exposure (professional or total exposure) or concussion history.

Emerging evidence from different football codes have increased urgency to investigate the chronic neurophysiological effects of repetitive sub-concussive head impacts (21). With evolving evidence that cumulative sub-concussive head impact exposure rather than concussion history may have a greater association on long term outcomes (4, 22). The aim of this exploratory study was to investigate, in a cohort of retired professional contact sport athletes, if changes assessed with TMS were associated with playing history exposure. Specifically, our question was whether a career exposure of contact sport participation (*total* career length from junior to retirement versus *professional* career span only) was associated with ageing neurophysiological changes as assessed by TMS. Secondary questions included if there were associations between TMS measures and participants’ reporting of fatigue and related symptoms affecting daily activities (23, 24), and the number of concussions experienced over their career.

## Materials & Methods

Retired male professional Australian football and rugby players (n=113; mean age [SD] 48.8 [9.6] years) were recruited for this study between 2018 to 2023 (see **table 1** for participant characteristics). Participants were excluded if they were retired less than five years or experienced any brain injury outside of contact sports participation (e.g., car or workplace accidents, fights or falls). Study participants reported no neurological, psychiatric, or neurodegenerative disease, sleep disorders (e.g., obstructive sleep apnoea), or current musculoskeletal injury, and provided written informed consent prior to data collection. Study protocols were approved by the University Human Research Ethics Committee (HEC18005).

**Table 1.**
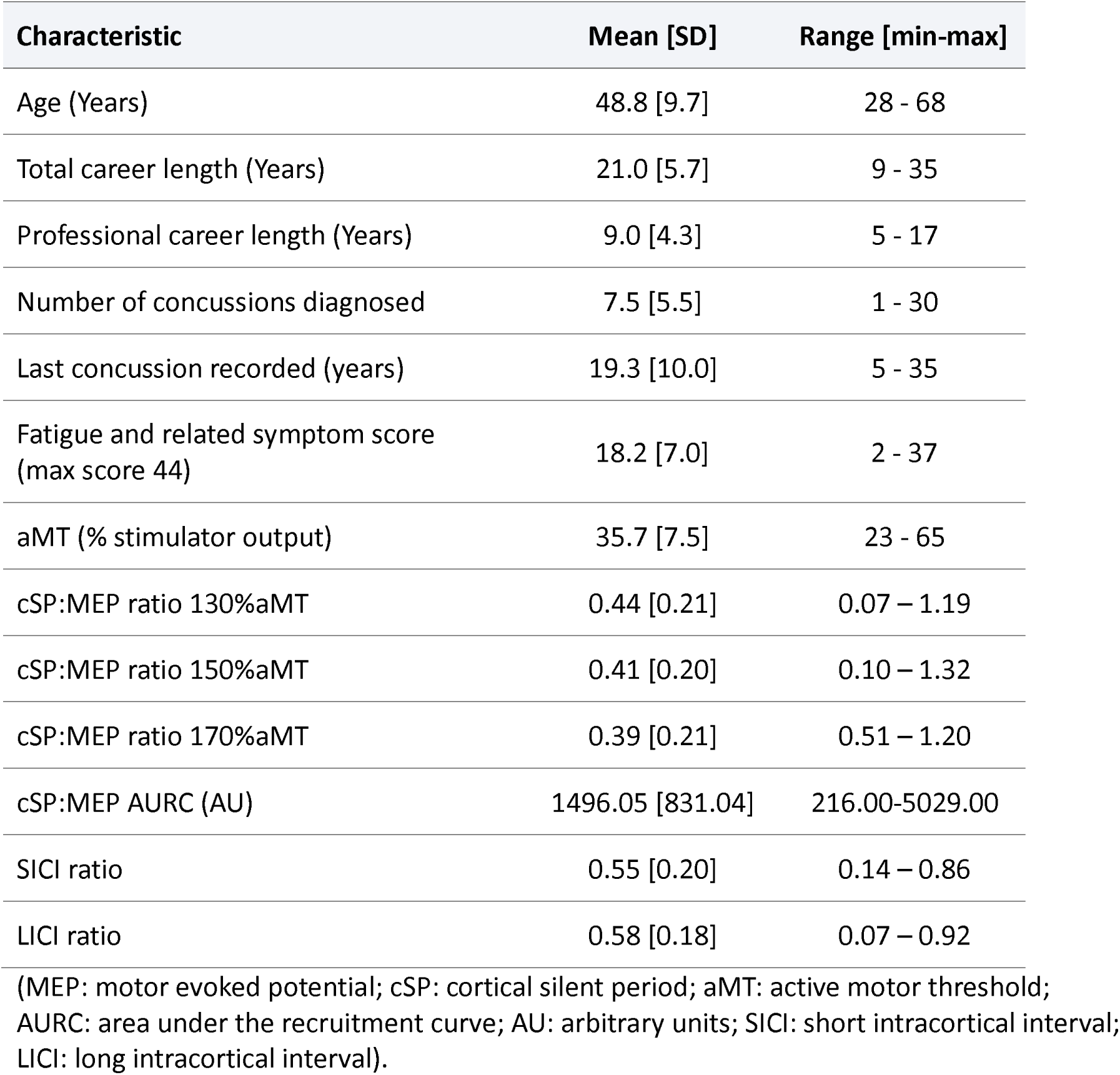
Participant characteristics (n=113, all male)

| Characteristic | Mean [SD] | Range [min-max] |
| --- | --- | --- |
| Age (Years) | 48.8 [9.7] | 28 - 68 |
| Total career length (Years) | 21.0 [5.7] | 9 - 35 |
| Professional career length (Years) | 9.0 [4.3] | 5 - 17 |
| Number of concussions diagnosed | 7.5 [5.5] | 1 - 30 |
| Last concussion recorded (years) | 19.3 [10.0] | 5 - 35 |
| Fatigue and related symptom score (max score 44) | 18.2 [7.0] | 2 - 37 |
| aMT (% stimulator output) | 35.7 [7.5] | 23 - 65 |
| cSP:MEP ratio 130%aMT | 0.44 [0.21] | 0.07 – 1.19 |
| cSP:MEP ratio 150%aMT | 0.41 [0.20] | 0.10 – 1.32 |
| cSP:MEP ratio 170%aMT | 0.39 [0.21] | 0.51 – 1.20 |
| cSP:MEP AURC (AU) | 1496.05 [831.04] | 216.00-5029.00 |
| SICI ratio | 0.55 [0.20] | 0.14 – 0.86 |
| LICI ratio | 0.58 [0.18] | 0.07 – 0.92 |

(MEP: motor evoked potential; cSP: cortical silent period; aMT: active motor threshold; AURC: area under the recruitment curve; AU: arbitrary units; SICI: short intracortical interval; LICI: long intracortical interval).

Participants completed the *fatigue and related symptom scores* questionnaire (23, 24) to quantify ongoing clinical concerns with daily activities. Players responded to 15 questions using a scale from ‘0’ (no concern) to ‘3’ (greatest concern) regarding levels of fatigue and difficulties they may encounter that impact on daily activities. The total score from the questionnaire was used to correlate to TMS parameters.

Motor evoked potentials (MEPs) from single and paired pulse TMS where quantified following stimulation over the contralateral motor cortex (9, 10, 20, 25). Surface electromyography (sEMG) recorded MEPs from 500 ms sweeps (100 ms pre-trigger, 400 ms post-trigger; PowerLab 4/35, ADInstruments, Australia) using two pre-gelled Ag/AgCl disposable surface electrodes (ADInstruments, Australia), placed over the first dorsal interosseous (FDI) muscle and a third ground electrode over the boney prominence of the fourth digit, of the participant’s dominant hand adhering to the Non-Invasive Assessment of Muscles (SENIAM) guidelines for sEMG (26).

TMS was delivered using a R30 stimulator with a C-B 60 coil (MagVenture, Denmark). For identification of the ‘optimal site’ (defined as the site with the largest observed MEP) on the motor cortex projecting to the FDI muscle, participants wore a snugly fitted cap (EasyCap, Germany), positioned with reference to the nasion-inion and interaural lines. The cap was marked with sites at 1 cm spacing in a latitude-longitude matrix to ensure reliable coil position throughout the testing protocol (27).

Identification of active motor threshold (aMT) was determined, during a low-level isometric contraction of the first index finder (targeting the FDI muscle) against a force transducer (ADInstruments, Australia) at 10% of maximal voluntary contraction (MVC). MVC was completed following a warm-up of graded increases in muscular contractions of the FDI (three contractions at intensities of 20%, 50% and 75% of their perceived MVC). Following the warm-up, participants then conducted two MVC trials of 3 s duration, each separated by 60 s of rest. Continuous verbal encouragement to push against the force transducer as hard as possible was provided. The maximum transducer value was recorded as the MVC. A third trial was cpmpleted if the difference between the first two trials exceeded 5% (28).

The aMT was identified by delivering TMS stimuli (5% of stimulator output steps, and in 1% steps closer to threshold) at intensities from a level well below expected participant’s threshold until an observable MEP of at 200 *µ*V and associated cortical silent period (cSP) could be measured in at least five of ten stimuli (29).

Participants abducted and controlled a static contraction of the contralateral index finger to target the FDI muscle (5% maximal voluntary contraction) during data collection. Stimulation was applied at 130%, 150% and 170% above aMT in order to explore stimulus-dependent corticomotor connectivity. Short intracortical inhibition (SICI) at 3ms interstimulus interval (ISI) was measured using a conditioning stimulus of 80% aMT and a test stimulus of 130% aMT, while long intracortical inhibition (LICI) was completed at 100 ms ISI using a suprathreshold conditioning and test stimulus at 130% aMT. Twenty single pulse stimuli (four sets of five pulses per set) were presented at random intervals, to avoid stimulus anticipation, four to eight seconds apart. For paired pulse (SICI and LICI), 20 stimuli were presented (four sets of five pulses per set) randomly six to 10 seconds apart. Between sets, a rest of 30 seconds was provided to reduce the possibility of muscular fatigue (30).

Single-pulse MEPs were measured as the amplitude of the waveform, and the cSP as the duration of silence in the EMG signal following the MEP waveform, and expressed as a cSP:MEP ratio, (9, 31) reflecting the balance between excitatory and inhibitory projections (lower ratios reflected reduced inhibition). Area under the recruitment curves (AURC) calculated MEP:cSP data at all stimulus intensities reflecting the input-output properties of the corticomotor pathway (32). SICI was calculated as a ratio of the paired-pulse MEP at 3ms to the single-pulse MEP at 130% aMT. LICI was calculated as a ratio of the suprathreshold test stimulus to the conditioning stimulus (33). As data was found to be non-normally distributed, relationships between variables were explored using Spearman’s *rho*. Data is presented as means (±SD) and alpha was set at 0.05, apart from cSP:MEP ratios where alpha was corrected as 0.017 due to comparisons at 130%, 150% and 170% aMT.

## Results

All participants recruited completed TMS testing with no adverse effects. Tables 1 and 2 show descriptive data and correlations respectively. All participants had reported starting to play their sport at 9.9 ± 2.4 years of age and retiring from their sport at 30.9 ± 4.5 years. All had reported at least one concussion in their career. For self-report fatigue and related symptoms, 72% of retired athletes scores were above the clinical cut-off as suggested by Johansson et al (23, 24).

**Table 2.** Correlations (rho) between TMS variables and concussions recorded, fatigue and related symptom scores, and career length (total and professional).

|  | Concussions<br>recorded | FRSS | Total career<br>length (years) | Professional career<br>length (years) |
| --- | --- | --- | --- | --- |
| aMT (% stimulator output) | -0.071<br>P=0.453 | -0.048<br>P=0.614 | 0.104<br>P=0.271 | -0.033<br>P=0.731 |
| MEP:cSP ratio 130%aMT | -0.174<br>P=0.066 | -0.319<br>P<0.001 | -0.321<br>P<0.001 | -0.106<br>P=0.265 |
| MEP:cSP ratio 150%aMT | -0.166<br>P=0.100 | -0.377<br>P<0.001 | -0.331<br>P<0.001 | -0.152<br>P=0.134 |
| MEP:cSP ratio 170%aMT | -0.157<br>P=0.118 | -0.274<br>P=0.006 | -0.331<br>P<0.001 | -0.148<br>P=0.142 |
| MEP:cSP AURC (AU) | -0.012<br>P=0.898 | -0.213<br>P=0.024 | -0.290<br>P=0.002 | -0.138<br>P=0.145 |
| SICI ratio | 0.158<br>P=0.095 | 0.144<br>P=0.129 | 0.263<br>P=0.005 | 0.164<br>P=0.076 |
| LICI ratio | 0.176<br>P=0.062 | 0.168<br>P=0.075 | 0.112<br>P=0.238 | 0.096<br>P=0.314 |

(FRSS: fatigue and related symptom score; MEP: motor evoked potential; cSP: cortical silent period; aMT: active motor threshold; AURC: area under the recruitment curve; SICI: short intracortical interval; LICI: long intracortical interval).

Significant correlations were found between total career length and cSP:MEP ratios, AURC, and SICI (*rho* -0.29 – -0.34, 0.25 respectively; *P*<0.05; table 2). Significant correlations were observed between symptom scores and cSP:MEP ratios and AURC (*rho* -0.21 – -0.38, *P*<0.05; table 2). No significant correlations were reported between the number of concussions recorded, or the length of the individual’s professional career on cSP:MEP ratios, SICI or LICI (table 2). Further no significant correlations were seen between symptoms scores with number of concussions, or career length (total or professional).

## Discussion

The aim of this exploratory study was to investigate in a cohort of ageing retired professional contact sport athletes the association of TMS neurophysiological outcomes with career exposure. While previous studies have reported changes associated with concussion history in retired contact sport athletes, (8–10) we found that impaired intracortical inhibition was significantly correlated with *total* career length and symptom reporting, but not with *professional* career length, or the number of recorded concussions. This is the first study showing pathophysiological findings associated with career experience, supporting recent neuropathological studies also reporting associations of exposure and risk of CTE (4, 22).

Previous TMS studies have reported increased cortical inhibition associated with healthy ageing (10, 34, 35) suggesting age-related changes in 1-aminobutyric acid (GABA) mediated cortical activity. This study, conversely, showed decreased cortical inhibition in a cohort of contact sport athletes, supporting previous concussion studies (9, 10, 20, 25). Of note, however, is that TMS data correlated to exposure rather than concussion history. Our results from this exploratory study provide further evidence of altered physiology in GABAergic receptor activity in cohorts exposed to repetitive neurotrauma. However, as TMS is an indirect measure, the mechanisms of repetitive sub-concussive trauma affecting GABAergic activity are postulated. In acute trauma studies, it has been shown that physical trauma alters depolarizing actions of GABA contributing to maladaptive signal transmission (36–39). It has been recently stated that repetitive sub-concussive head impact research using TMS is needed (21). The data in our TMS study suggests that repetitive and regular sub-concussive impacts experienced by athletes in contact sports over multiple seasons are cumulative, and consequently contribute to maladaptive long-term potentiation (LTP) changes altering pre/post synaptic processing (10, 40). While we appreciate that the majority of LTP work stems from experimental stimulation studies (40), LTP has also been shown not only in neurological and psychiatric conditions but also with interventional methods such as exercise (41). It is plausible that our findings reflect, in this cohort, the experiences of individuals who have experienced tens of thousands of sub-concussive impacts (4) over their playing lifetime. Further research extending on this work, using spatio-temporal techniques such as magnetoencephalography to illustrate multi-region functional LTP connectivity, could confirm these initial findings.

Conversely, the finding of no correlations between the number of concussions and TMS data was surprising. Previous studies (8, 9, 19, 20, 25) investigated cortical physiology changes in light of concussion history only. While a majority of these studies found differences between athletes and age-matched controls (8, 9, 20, 25), what the data from our study suggests is that previous findings could have been ascribed to career exposure rather than concussion history. Future studies investigating long term outcomes in these cohorts should record career history that includes when participant’s started playing contact sports and when they completely retired from their sport further to their concussion history.

A secondary finding was the significant correlation of higher symptom scores with decreased cortical inhibition. Although self-reported in nature , high reliability of the scale (Chronbach’s alpha of 0.94 (23)) gives us confidence that participants did not exaggerate their responses, as we would not have seen a significant correlation of symptom scores to TMS data. Moreover, our data concurs with previous studies showing differences in TMS excitability and inhibition between groups (described as ‘symptomatic’ and ‘asymptomatic’) (9) separated by the cut-off score as recommended by Johansson and colleagues (23, 24). We are also aware of the limitations of self-report of concussion history, but aimed to only record concussions where the player recalled being assessed by the club doctor and missed the following week of competition (25).

Another limitation of this study is that TMS with sEMG is a measure of corticomotor physiology. Although cSP, SICI, and LICI evoked potentials represents intracortical networks, we caution on generalizing these findings to non-motor regions. Further, our SICI and LICI protocols were limited to only 3 ms and 100 ms ISIs respectively. Future studies should aim to include paired pulse TMS with electroencephalography, previously reported in those with persistent post-concussion symptoms (34), and a panel of ISIs for SICI and LICI, as well as utilizing intracortical facilitation, which has been undertaken in recent dementia studies (16, 18).

Emerging evidence is suggesting that exposure to repetitive neurotrauma is the main determinant of risk of CTE (4, 22). However, CTE can only be diagnosed post-mortem, therefore a major challenge is to aim to identify ageing individuals at a heightened risk for neurological decline and possible dementia as early as possible (14). While a TES clinical criteria is being developed, as yet there is no agreement on a biomarker as part of the clinical criteria (6). As TMS has previously shown to differentiate between early stage Alzheimer’s disease and frontotemporal dementia (18), there are opportunities to utilise TMS as a biomarker towards clinical diagnostic utility for TES (14).

In conclusion our findings, demonstrating abnormal cortical physiology associated with total career exposure, suggest that TMS evoked potentials would be a suitable prodromal biomarker to complement clinical assessment and neuropsychological components in developing diagnostic criteria for TES. We call to progress this research further, so that objective measures can assist diagnostics for those with a history of neurotrauma suspected of CTE.

## Data availability

The data generated during and/or analysed during the current study are not publicly available due to potential ability to identify professional athletes based on exposure data. However, these data are available from the corresponding author upon reasonable request and with relevant University Human Research Ethics approval.

